# Safety and feasibility of an FES-Cycling based neuromuscular evaluation method in mechanically ventilated patients

**DOI:** 10.1101/2022.04.11.22273563

**Authors:** Thainá de Gomes Figueiredo, Murillo Frazão, Luís Augusto Werlang, Cássio Azevedo, Adelar Kunz, Maikel Peltz, Veridiana Câmara Furtado, Edgar Brito de Souza, Júlio Francisco de Moura, Rosane Maria Silva, Dário Celestino Sobral Filho

## Abstract

**Purpose:** FES-Cycling can be used to assess neuromuscular performance, however the safety and feasibility of this evaluation method has never been investigated.

**Materials and methods:** an observational prospective study was carried out. The FES-Cycling equipment was set in the evaluation mode. For safety determination, hemodynamic parameters and peripheral oxygen saturation were measured before and immediately after the evaluation protocol, as well as venous oxygen saturation and blood lactate. The creatine phosphokinase level (CPK) was measured before and 24, 48 and 72 hours after the test. The time spent to carry out the entire evaluation protocol and the number of patients with visible muscle contraction were recorded to assess feasibility.

**Results:** Heart rate, systolic and diastolic blood pressure did not change after protocol evaluation, as well as peripheral and venous oxygen saturation (p > 0.05). Moreover, blood lactate did not change (p > 0.05). CPK did not change up to 72 hours after the test (p > 0.05). The time for evaluation was 11.3 (SD = 1.1) minutes. Furthermore, 75% of the patients presented very visible muscle contraction, 25% of the patients presented barely visible and no patients presented non-visible muscle contraction.

**Conclusions:** FES-Cycling based neuromuscular evaluation method is safe and feasible.

## 1. Introduction

New technologies, multidisciplinary therapeutic approaches and developments in managing critically ill patients significantly increased survival rates in intensive care units (ICU). At the same time, several sequelae emerged in ICU survivors, such as intensive care unit-acquired weakness (ICU-AW)(1). The most common neuromuscular disorders found in ICU patients are myopathy, polyneuropathy and a combination of these two disorders, polyneuromyopathy(2).

ICU-AW is characterized by a decrease in the neuromuscular excitability and generalized muscle weakness(3). The development of ICU-AW is associated with high functional impairment and mortality even after years of hospital discharge(4)(5). Several diagnostic methods have been used to detect ICU-AW. They range from simple strength measurement (Medical Research Council score and dynamometry)(6) to non-volitional neuromuscular evaluations (evocated peak torque, chronaxie determination, electroneuromyography, etc.)(7)(8)(3).

Functional electrical stimulation associated to cyclergometry (FES-cycling) has gained popularity in all situations in which patients cannot actively move their legs. The concept is to promote cyclergometry exercise induced by depolarization of the motoneuron and consequently all the physiological stages of muscular contraction. It uses computer-driven electrical pulses delivered by transcutaneous electrodes, independently promoting muscle contractions on functionality of the physiological pathway. This muscle contraction must be synchronized, harmonic and adequate to promote cycling movement(9).

The FES-Cycling technology can be used to assess neuromuscular performance. The equipment objectively provides an electrical stimulus and measures the muscle’s mechanical response to power output and torque(10). Additionally, the technology can also measure the stimulation cost (total electrical charge delivery rate per watt of power output)(11), providing information on the neuromuscular physiological pathway.

The safety and feasibility of the FES-Cycling based neuromuscular evaluation method has never been investigated in critical ill patients. Thus, the aim of this study was to evaluate the safety and feasibility of this evaluation method in critically ill patients undergoing mechanical ventilation. Our hypothesis was that this evaluation method would be feasible, safe, and not promote a disturbance in vital signs, in the relationship between supply and oxygen consumption, or cause neuromuscular harm.

## 2. Material and methods

### 2.1 Study design

An observational prospective study was carried out from December 2021 to February 2022 in the ICU of a cardiology reference hospital in Brazil. The protocol was approved by local ethics committee in compliance with the Declaration of Helsinki (opinion number 5.069.827/21, CAAE: 50202821.1.0000.9030) and was registered in the Brazilian Clinical Trial Registration Platform (Number: RBR-10gyv7wn). Those legally responsible for the patients signed a free and informed consent form before the study. Patients were consecutively admitted and the evaluation protocol was applied in each patient. The study included individuals older than 18 years, both genders, quadriparetic and mechanically ventilated. Patients with hemodynamic instability (mean arterial pressure < 65 or > 110 mmHg) or patients with skin or musculoskeletal lesions which prevented performing FES-Cycling were excluded.

### 2.2 Evaluation protocol

The patients were attached to the FES-Cycling equipment (Figure 1) (MOBITRONICS^®^, INBRAMED, Brazil). Equipment height and distance and leg support positions were individually adjusted for the best biomechanical fit. The skin was cleaned and trichotomy was performed if necessary before electrode placement. Self-adhesive electrodes were placed bilaterally on the belly of the quadriceps (vastus lateralis and vastus medialis), hamstrings and tibialis anterior muscles, and then plugged to the electrical stimulation device cables. Eight electrical stimulation channels were used. FES was set with the same pulse width and intensity in all eight channels (500μs pulse width for 50-100mA intensity; 600μs pulse width for 101-130mA intensity; 700μs pulse width for 131-160mA intensity; 800μs pulse width for 161-190mA intensity; 900μs pulse width for 191-220mA intensity and 1000μs pulse width for 221-250mA intensity). One channel was activated for one second to detect the quality of muscle contraction before the evaluation started. FES parameters of pulse width and intensity were set to promote the highest visible muscular contraction without pain.

**Figure 1:**
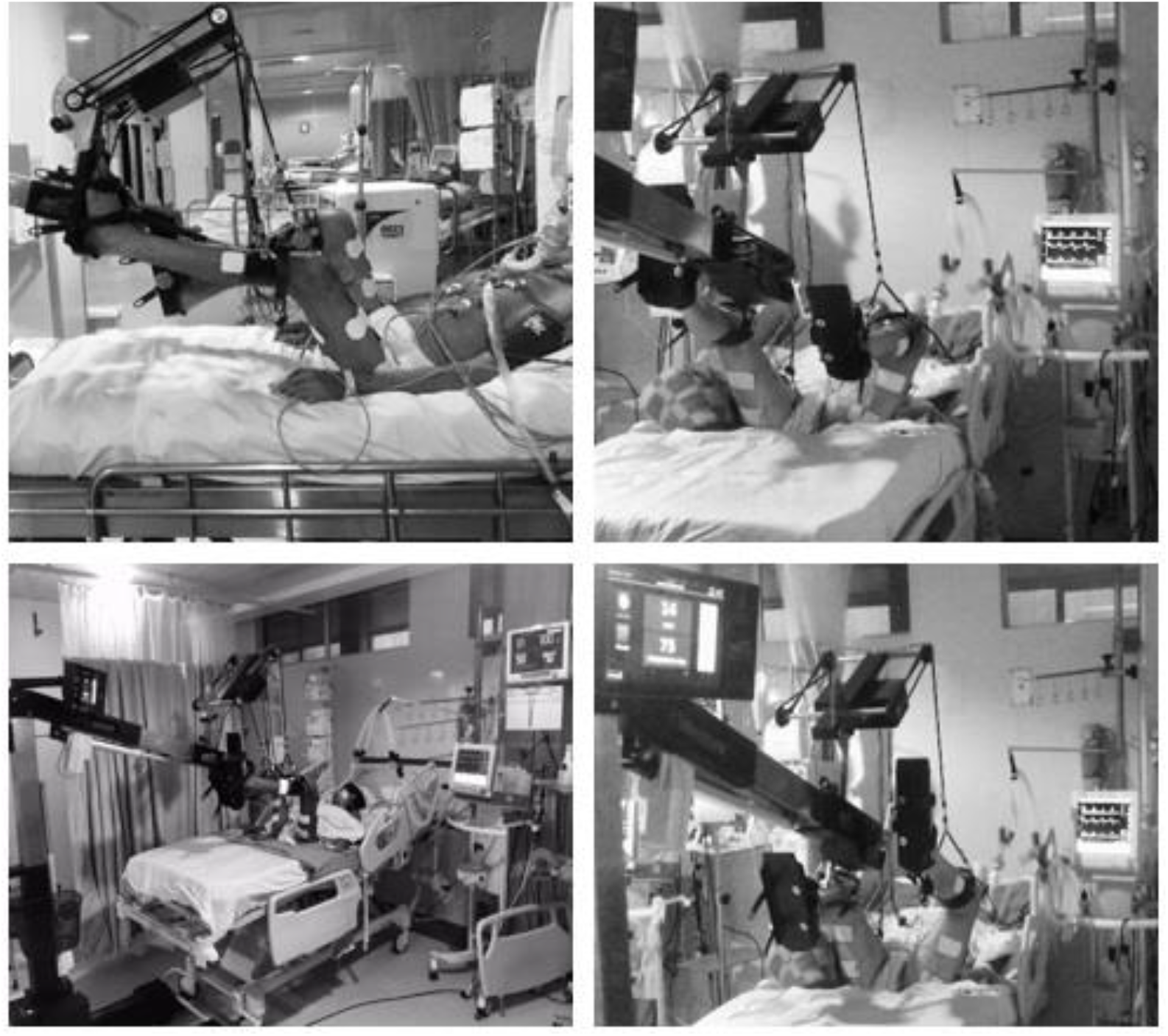
Patients attached to the FES-Cycling equipment.

Pain was evaluated in conscious patients by self-report. The patients were asked to answer ‘‘yes or no’’ by nodding their head to the question: ‘‘Does it hurt?’’. Pain in unconscious patients was evaluated by Critical-Care Pain Observation Tool(12), with a cut-off point ≥ 2 for pain. FES was triggered (ON) and stopped (OFF) by pedal position. The equipment has a sensor to detect 360° pedal position. FES trigger/stop was set according to physiological joints (hip, knee and ankle) positions during cycling movement. The equipment was set in the evaluation mode (Power Test^®^). It performed a combination of different cyclergometry cadences (10, 15 and 20 rotation per minute) and electrical stimulation frequencies (50, 75 and 100Hz). The patients performed 10 cycling movements in each combination (90 cycling movements in total). The patients did not perform any voluntary effort. All the work was performed by the FES-Cycling equipment. The equipment recorded the maximum power output and torque reached, in addition to stimulation cost.

### 2.3 Safety and feasibility assessment

The safety and feasibility assessment protocol was based on the one previously described by Silva et al.(13). Patients were evaluated for 3 consecutive days after testing to determine the safety. To do so, 4 mL blood samples were collected daily from the central venous access. Hemodynamic parameters (blood pressure and heart rate) and peripheral oxygen saturation were measured before and immediately after the evaluation protocol (IPM-9800, Mindray, China), as well as venous oxygen saturation and blood lactate (ABL 800 flex, Radiometer, Denmark). The creatine phosphokinase level in the blood (CPK) was measured before (baseline) and 24, 48 and 72 hours after the test (Vitros XT 7600, Ortho Clinical Diagnostics, USA). Baseline high and low CPK levels were established by the reference values for healthy subjects provided by the manufacturer (130 IU/L for female and 170 IU/L for male).

The time spent to carry out the entire evaluation protocol was recorded to assess feasibility. The total time was divided into: 1) time to prepare patient (attach patient in the equipment, place the electrodes, plug the cables, set pulse width and intensity parameters); 2) time to set angles for trigger/stop; 3) time to test execution (execution of the 90 cycling movements); and 4) time for equipment release (unplug the cables, remove the electrodes and take off the equipment). The number of patients with very visible, barely visible or non-visible muscle contraction was also recorded.

### 2.4 Statistical analysis

Data normality was verified using the Shapiro-Wilk test. Data are presented as means ± standard deviations or as medians and interquartile ranges (according to data normality) and percentages. Differences in hemodynamic parameters, peripheral oxygen saturation, venous oxygen saturation and blood lactate were evaluated by paired t-test or Wilcoxon test (according to data normality). Differences in the CPK level were evaluated by the Friedman test with Dunn’s multiple comparisons test. The post hoc achieved power of effect size was also computed(14). The effect size convention for hemodynamic parameters, peripheral oxygen saturation, venous oxygen saturation and blood lactate was: trivial < 0.2; small > 0.2; medium > 0.5 and large > 0.8 (t-test family). The effect size convention for CPK level was: small > 0.1, medium > 0.25 and large > 0.40 (F-test family). A statistically significant value of p < 0.05 was set for all analyses. GraphPad Prism 7.0 and GPower 3.0.10 software programs were used.

## 3. Results

A total of 26 patients were enrolled in the study, but 6 were excluded. The characteristics of the patients are presented in Table 1. Pulse width, intensity, power output, torque and stimulation cost are presented in Table 2.

**Table 1.** Patients characteristics.

| Variable | Value |
| --- | --- |
| *Age (years)* | 61 ± 20 |
| *Sex (male/female)* | 13/7 |
| *ICU stay (days)* | 6.0 (2.0 – 8.0) |
| *Mechanical ventilation (days)* | 4.5 (2.0 – 8.0) |
| *SAPS III* | 71 ± 10 |
| *24h water balance (mL)* | 1275 ± 1562 |
| *Glucose (mg/dL)* | 178 ± 79 |
| *Vasoactive drugs use (n,%)* | 14 (70) |
| *Sedation use (n,%)* | 17 (85) |
| *Corticoids use (n,%)* | 6 (30) |
| *Lower limbs edema (n,%)* | 10 (50) |
| *Sepsis (n,%)* | 19 (95) |
| *Main diagnosis* |  |
| *Acute myocardial infarction (n,%)* | 4 (20) |
| *Heart failure (n,%)* | 2 (10) |
| *Shock (n,%)* | 4 (20) |
| *Arrhythmia (n,%)* | 7 (35) |
| *Myocarditis (n,%)* | 1 (5) |
| *Acute pulmonary edema (n,%)* | 1 (5) |
| *Acute respiratory failure (n,%)* | 1 (5) |

ICU: Intensive care unit; SAPS III: Simplified Acute Physiology Score III. Mean ± standard deviation. Median (interquartile range).

**Table 2.** Pulse width, intensity, power output, torque and stimulation cost.

| Variable | Value |
| --- | --- |
| *Pulse width (μs)* | 600 (500 – 800) |
| *Intensity (mA)* | 124 ± 46 |
| *Power output (watts)* | 2.9 (1.8 – 4.1) |
| *Torque (N/m)* | 1.9 (1.1 – 4.3) |
| *Stimulation cost (μC/w)* | 27,666 (15,371 – 57,975) |

μs: microseconds, mA: miliamper, Nm: newton per meter, μC/w: microcoulombs per watt. Median (interquartile range). Mean ± standard deviation.

### 3.1 Safety

Heart rate = 91 ± 23 vs 94 ± 23bpm (p = 0.0837); systolic blood pressure = 122 ± 19 vs 124 ± 19mmHg (p = 0.4261) and diastolic blood pressure = 68 ± 13 vs 70 ± 15mmHg (p = 0.3462) did not change after protocol evaluation (trivial effect size) (Figure 2). Peripheral oxygen saturation = 98 (96-99) vs 98 (95-99)% (p = 0.6353) and venous oxygen saturation = 71 ± 14 vs 69 ± 14% (p = 0.1317) also did not change after protocol evaluation (trivial effect size) (Figure 3). Moreover, blood lactate = 1.48 ± 0.65 vs 1.53 ± 0.71mmol/L (p = 0.2320) did not change after protocol evaluation (trivial effect size) (Figure 4). All patients CPK level = 99 (59 – 422) vs 125 (66 – 674) (p = 0.2799) vs 161 (66 – 352) (p > 0.999) vs 100 (33 – 409) (p = 0.5901) did not change up to 72 hours after the test (with small effect size) (Figure 5). High CPK level patients = 637 (315 – 1403) vs 707 (375 – 1324) (p = 0.4419) vs 463 (286 – 1049) (p = 0.9019) vs 452 (315 – 1199) (p > 0.999) did not change up to 72 hours after protocol evaluation (small effect size) (Figure 5). Low CPK level patients = 66 (46 – 99) vs 73 (50 – 93) (p = 0.9653) vs 79 (39 – 181) (p > 0.999) vs 45 (29 – 107) (p = 0.9653) did not change up to 72 hours after the test (small effect size) (Figure 5).

**Figure 2:**
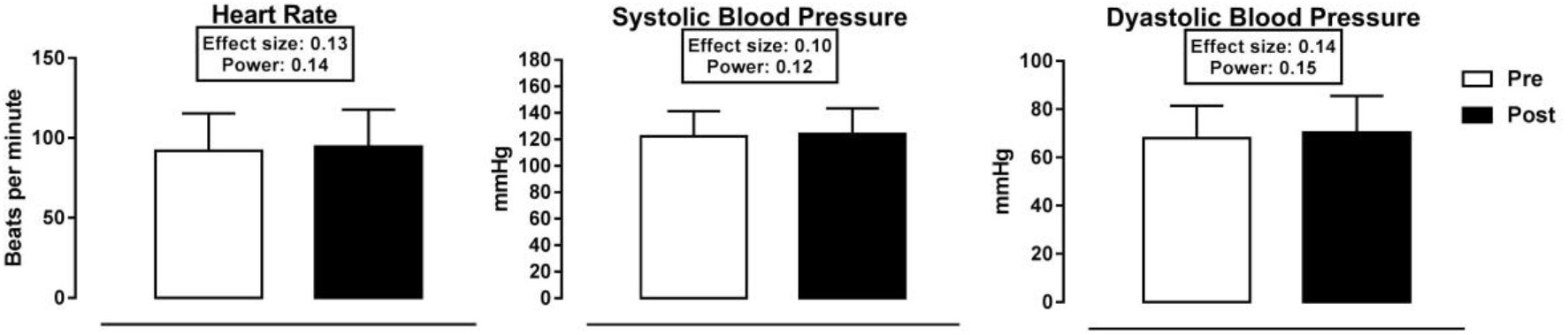
Hemodynamic parameters.

**Figure 3:**
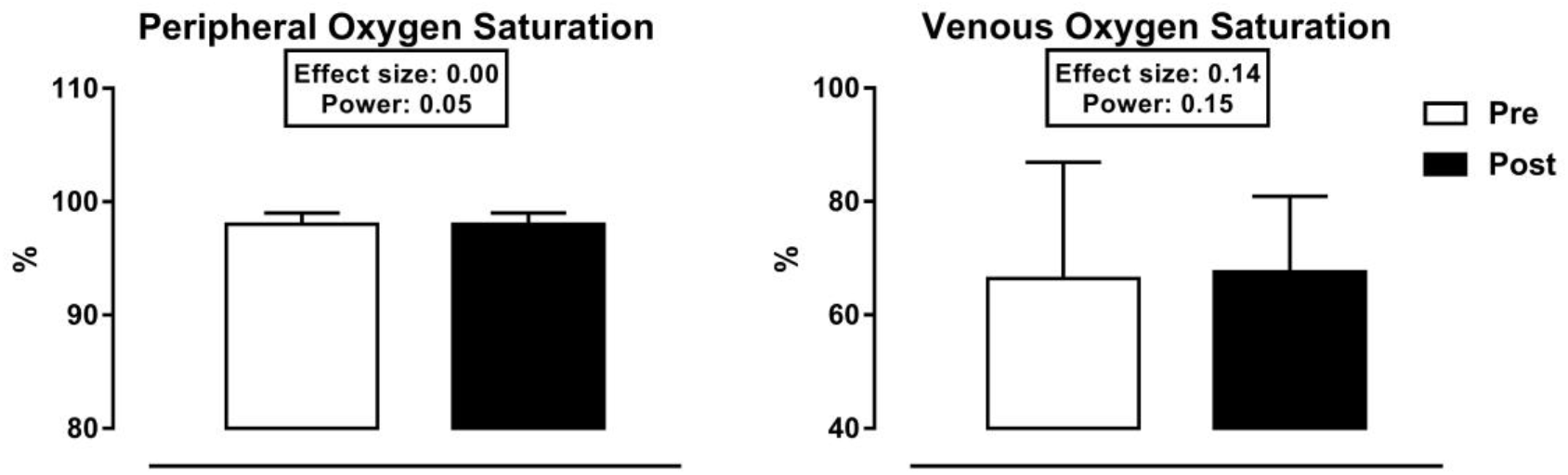
Peripheral and venous oxygen saturation.

**Figure 4:**
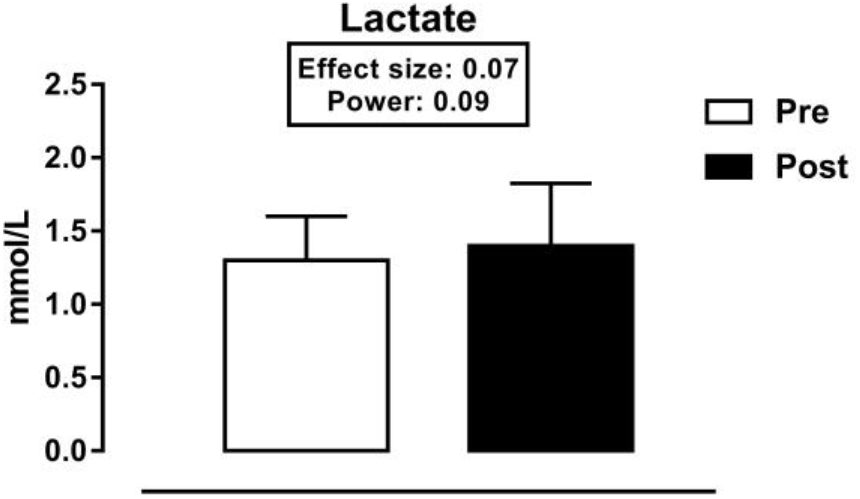
Lactate level.

**Figure 5:**
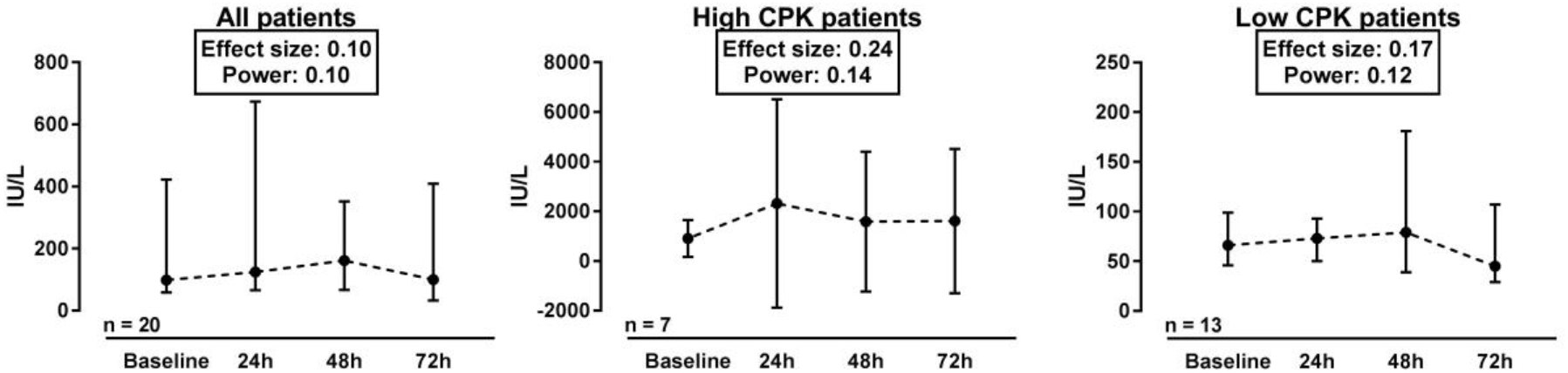
Creatine phosphokinase level.

### 3.2 Feasibility

The total time was 11.3 ± 1.1 minutes. The time to prepare the patient was 5.0 ± 1.1 minutes (43% of total time); time to set pulse width and intensity parameters and angles for trigger/stop was 0.6 (0.5 – 0.7) minutes (7% of total time); time for test execution was 4.4 ± 0.1 (39% of total time) minutes; and time for equipment release was 1.3 ± 0.3 minutes (11% of total time) (Figure 6). Furthermore, 15 patients (75%) presented very visible muscle contraction, 5 patients (25%) presented barely visible and no (0%) patients presented non-visible muscle contraction (Figure 7).

**Figure 6:**
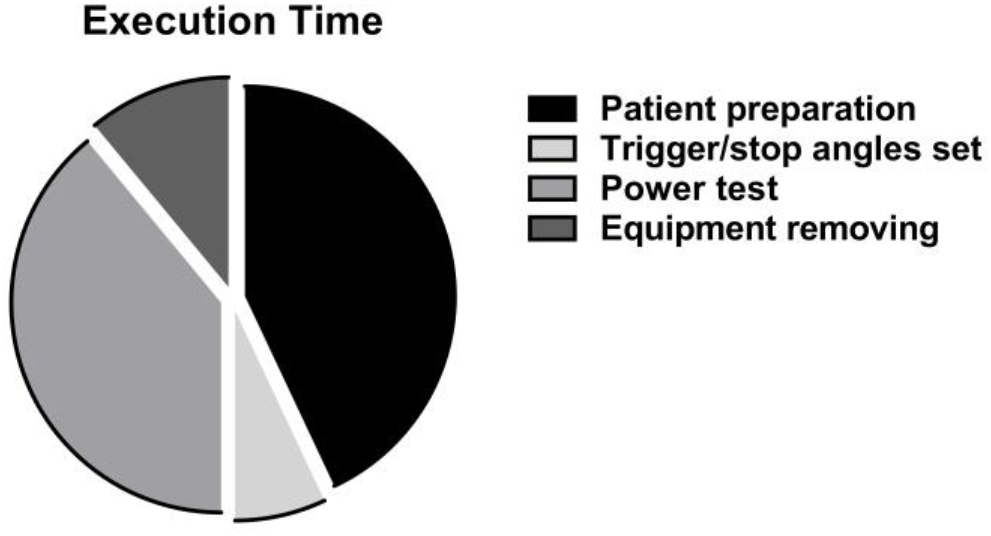
Execution time.

**Figure 7:**
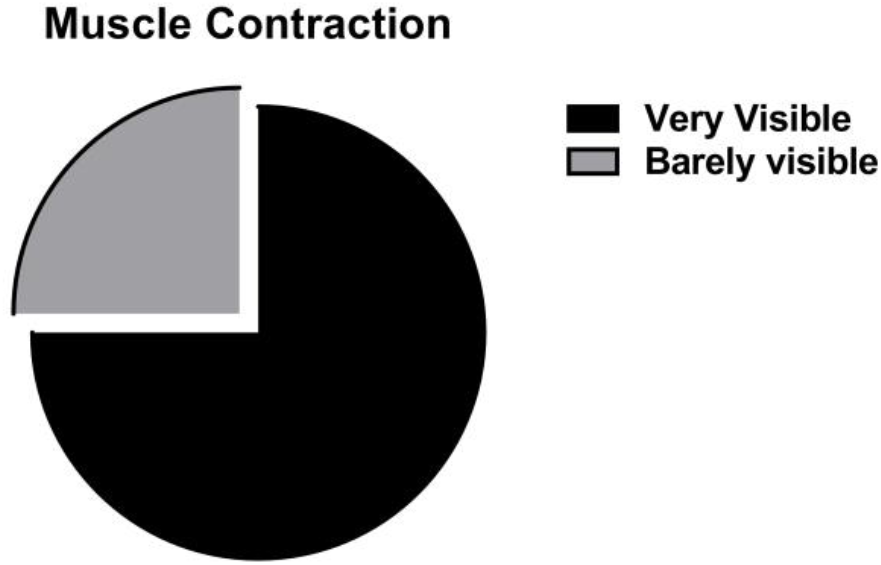
Muscle contraction visibility.

## 4. Discussion

To summarize our results we can state that: 1) The FES-Cycling based neuromuscular evaluation method is safe and feasible; 2) Hemodynamic parameters, peripheral oxygen saturation, venous oxygen saturation and blood lactate did not change after the evaluation protocol; 3) CPK did not increase up to 72h after evaluation protocol; 4) The FES-Cycling based neuromuscular evaluation method only needed a few minutes to be performed; and 5) the majority of the patients presented very visible muscle contraction.

Non-volitional force measurements have been reported in mechanically ventilated patients. It generally consists of a force transducer and an adjustable platform combined with supramaximal stimuli applied either over a motor nerve or a muscle belly(15). Non-commercial ergometers have been developed which enable recording evoked force in ICU patients in different muscle groups: adductor pollicis(16), ankle dorsiflexors(17) and quadriceps(7). However, it is unclear whether and to what extent force measurements on a single muscle group may be representative for generalized muscle weakness.

This is the first study to report the use of a cyclergometer to assess neuromuscular function. The main advantages of FES-cycling based method are the possibility of measuring the simultaneous responses of a large group of muscles, constituting a dynamic evaluation using functional movement, not requiring high specialized expertise, and can be performed in the very early phase of ICU admission.

There were no adverse events during the protocol evaluation in the present study. The safety of FES-Cycling in critically ill patients has already been demonstrated in previous studies. In accordance with our findings, Parry et al.(18) demonstrated no major adverse events during FES-Cycling in mechanically ventilated patients, with only one minor adverse event occurring in the 30-minute post training period. This subject had a transient peripheral oxygen desaturation to 86% for greater than 1 minute, requiring a temporary increase in fraction of inspired oxygen from 0.4 to 0.6 for 1 hour.

Medrinal et al.(19) demonstrated heart rate and blood pressure increase during FES-Cycling in mechanically ventilated patients, however no adverse events were reported. Our data show that hemodynamic parameters return to baseline values just after exercise cessation. Godja et al.(20) demonstrated a mild elevation of arterial lactate and reduced venous oxygen saturation during FES-Cycling in healthy subjects. Our data show no significant change in these variables. This discrepancy is probably due to the time and place of blood sample collection. Arterial lactate increases during exercise as a result of isocapnic buffering of lactic acid produced during glycolytic muscle fiber contraction(21). We collected a venous lactate sample after the exercise. During FES-Cycling, arterial-mixed venous oxygen content difference(9) and oxygen extraction(19) are increased, reducing venous oxygen saturation. As we collected the blood sample after exercise, there was no oxygen extraction increase at this moment.

Skeletal muscle damage can be indirectly assessed by blood CPK levels. The FES-Cycling based neuromuscular evaluation method promoted no muscle damage. Muscle tissue may be damaged following intense prolonged activation as a consequence of both metabolic and mechanical factors. Indeed, rhabdomyolysis may result from direct and indirect damage to the muscle membrane, and may lead to leakage of intracellular muscle components into the extracellular fluid(22). In cases of electrical stimulation-induced muscle damage, creatine kinase levels increase in the first day after stimulation(23). In the present study, no significant variation in CPK levels up to 72h after evaluation protocol was found, even in high baseline CPK level patients. The three highest baseline CPK level patients (>1,200 IU/L) had cardiac arrest up to 96h prior to the evaluation. Creatine kinase elevation is a common finding following successful cardiopulmonary resuscitation after cardiac arrest and this elevation is related to both physical as well as electrical injury (defibrillation) sustained during cardiopulmonary resuscitation(24)(25).

The short time spent to perform the entire exam is important for adherence to the evaluation method. As the FES-Cycling based neuromuscular evaluation method only requires a few minutes, it can be easily performed in a large number of patients in the same day. It can also be done as a routine evaluation before each therapeutic FES-Cycling session, providing neuromuscular monitoring during the ICU stay.

All patients presented visible muscle contraction during evaluation. The majority of the patients presented very visible muscle contraction. Segers et al.(26) demonstrated that critically ill patients with sepsis, edema, or receiving vasopressors are less likely to respond to electrical stimulation with adequate muscle contraction. We had adequate muscle contraction in the present study even in these situations. A possible reason for this was the FES parameters set, with 600μs median pulse width and 124mA mean intensity. Another possible reason was the cycling movement that changed muscle length and position, improving motor unit recruitment.

This study has some limitations. First, the sample size was not calculated, and a convenience sample of consecutive patients was used. Second, cardiac patients compose almost the entire sample due to the hospital profile.

## 5. Conclusions

FES-Cycling based neuromuscular evaluation method is safe and feasible. Hemodynamic parameters, peripheral oxygen saturation, venous oxygen saturation and blood lactate did not change after the evaluation protocol. Muscle damage markers did not increase up to 72h after the evaluation protocol. The entire evaluation only needed a few minutes to be performed and the majority of the patients presented very visible muscle contraction.

## Data Availability

All data produced in the present work are contained in the manuscript

## Abbreviations

ICU: intensive care unit
ICU-AW: intensive care unit-acquired weakness
FES-Cycling: functional electrical stimulation associated to cyclergometry
CPK: creatine phosphokinase

## CRediT authorship contribution statement

**Thainá de Gomes Figueiredo**: Conceptualization, Methodology, Data cutration, Formal analysis, Investigation, Writing – original draft, Writing – review & editing. **Murillo Frazão**: Conceptualization, Methodology, Formal analysis, Investigation, Writing – original draft, Writing – review & editing. **Luís Augusto Werlang, Cássio Azevedo, Adelar Kunz and Maikel Peltz**: Investigation, Writing – original draft, Writing – review & editing. **Veridiana Câmara Furtado, Edgar Brito de Souza Júnior, Júlio Francisco de Moura Júnior and Rosane Maria Silva**: Methodology, Data curation, Investigation, Writing – review & editing. **Dário Celestino Sobral Filho**: Conceptualization, Methodology, Formal analysis, Investigation, Writing – original draft, Writing – review & editing.

## References

1. Kress JP, Hall JB. ICU-Acquired Weakness and Recovery from Critical Illness. N Engl J Med. 2014;970(17):1626–35.

2. Lacomis D. Neuromuscular Disorders in Critically Ill Patients : Review and Update. J Clin Neuromusc Dis. 2011;12(4):197–218.

3. Lacomis D. Electrophysiology of Neuromuscular Disorders in Critical Illness. Muscle Nerve. 2013;47:452–63.

4. Herridge MS, Tansey CM, Matté A, Tomlinson G, Diaz-Granados N, Cooper A, Guest CB, Mazer CD, Mehta S, Stewart TE, Kudlow P, Cook D, Slutsky AS CA. Functional disability 5 years after acute respiratory distress syndrome. N Engl J Med. 2011;764(14):1293–304.

5. Dinglas VD, Friedman LA, Colantuoni E, Mendez-tellez PA, Shanholtz CB, Ciesla ND. Muscle Weakness and 5-Year Survival in Acute Respiratory Distress Syndrome Survivors. Crit Care Med. 2018;45(3):446–53.

6. Vanpee G, Hermans G, Segers J, Gosselink R. Assessment of Limb Muscle Strength in Critically Patients: A Systematic Review. Crit Care Med. 2014;42(3):701–11.

7. Laghi F, Khan N, Schnell T, Alexonis D, Hammond K, Shaikh H, et al. New device for nonvolitional evaluation of quadriceps force in ventilated patients. Muscle Nerve. 2019;57(5):784–91.

8. Tatjana Paternostro-Sluga, Othmar Schuhfried, Gerda Vacariu, Thomas Lang VF-M. Chronaxie and accommodation index in the diagnosis of muscle denervation. Am J Phys Med Rehabil. 2002;81(4l):253–60.

9. Frazão M, Augusto L, Azevedo C, Kunz A. Metabolic, ventilatory and cardiovascular responses to FES-cycling : A comparison to NMES and passive cycling. Technol Heal Care. 2021;doi: 10.3233/THC-213220.

10. Gföhler M, Lugner P. Dynamic Simulation of FES-Cycling : Influence of Individual Parameters. IEEE Trans Neural Syst Rehabil Eng. 2004;12(4):398–405.

11. Hunt KJ, Fang J, Saengsuwan J, Grob M, Laubacher M. On the efficiency of FES cycling : A framework and systematic review. Technol Heal Care. 2012;20(5):395–422.

12. Céline Gélinas CJ. Pain Assessment in the Critically Ill Ventilated Adult : Validation of the Critical-Care Pain Observation Tool and Physiologic Indicators. Clin J Pain. 2007;23(6):497–505.

13. Paulo Eugênio Silva, Nicolas Babault, João Batista Mazullo, Tamires Pereira de Oliveira, Bárbara Letícia Lemos, Vitor Oliveira Carvalho JLQD. Safety and feasibility of a neuromuscular electrical stimulation chronaxie-based protocol in critical ill patients: A prospective observational study. J Crit Care. 2017;37:141–8.

14. Cohen J. A power prime. Psychol Bull. 1992;112(1):155–9.

15. Kennouche D, Luneau E, Lapole T, Morel J, Millet GY. Bedside voluntary and evoked forces evaluation in intensive care unit patients : a narrative review. Crit Care. 2021;25(1):157.

16. Finn PJ, Plank LD, Clark MA, Connolly AB, Hill GL. Assessment of Involuntary Muscle Function in Patients After Critical Injury or Severe Sepsis. J Parenter Enter Nutr. 1996;20(5):332–7.

17. HF Ginz, PA Iaizzo, Albert Urwyler HP. Use of non-invasive-stimulated muscle force assessment in long-term critically ill patients: a future standard in the intensive care unit? Acta Anaesthesiol Scand. 2008;52(1):20–7.

18. Parry SM, Berney S, Warrillow S, El-ansary D, Bryant AL, Hart N, et al. Functional electrical stimulation with cycling in the critically ill : A pilot case-matched control study. J Crit Care. 2014;29(4):695.e1-7.

19. Medrinal C, Combret Y, Prieur G, Quesada AR, Bonnevie T, Gravier FE, et al. Comparison of exercise intensity during four early rehabilitation techniques in sedated and ventilated patients in ICU : a randomised cross-over trial. Crit Care. 2018;22(1):1–8.

20. Gojda J, Waldauf P, Hrušková N, Blahutová B, Krajčová A, Urban T, Tůma P, Řasová KDF. Lactate production without hypoxia in skeletal muscle during electrical cycling : Crossover study of femoral venous-arterial differences in healthy volunteers. PLoS One. 2019;14(3):1–15.

21. Wasserman K WB. Excercise physiology in health and disease. Am Rev Respir Dis. 1975;112(2):219–49.

22. Brancaccio P, Lippi G, Ematochimica UOD, Ospedaliero-A. Biochemical markers of muscular damage. Clin Chem Lab Med. 2010;48(6):757–67.

23. Alexandre Fouré, Kazunori Nosaka, Jennifer Wegrzyk, Guillaume Duhamel, Arnaud Le Troter, Hélène Boudinet, Jean-Pierre Mattei, Christophe Vilmen, Marc Jubeau, David Bendahan JG. Time Course of Central and Peripheral Alterations after Isometric Neuromuscular Electrical Stimulation-Induced Muscle Damage. PLoS One. 2014;9(9):e107298.

24. Miillner M, Sterz F, Domanovits H, Laggner AN. Creatine kinase and creatine kinase-MB release after nontraumatic cardiac arrest. Tha Am J Cardiol. 1996;77(8):581–5.

25. Joseph Mattana PCS. Determinants of Elevated Creatine Kinase Activity and Creatine Kinase MB-Fraction following Cardiopulmonary Resuscitation. Chest. 1990;101(5):1386–92.

26. Segers J, Hermans G, Bruyninckx F, Meyfroidt G, Langer D, Gosselink R. Feasibility of neuromuscular electrical stimulation in critically ill patients. J Crit Care. 2014;29(6):1082–8.

